# The health effects of Chunga Dumpsite on surrounding communities in Lusaka, Zambia

**DOI:** 10.1101/2021.12.21.21268110

**Authors:** Wilson Chibwe, Aaron Mbewe, Alice Ngoma Hazemba

## Abstract

**Background:** Dumpsites in low and middle income countries (LMICs) are commonly used for solid waste disposal as opposed to landfills. However, these are naturally unsanitary hence provide ideal conditions for breeding of disease transmitting agents and become predisposing factors for spread of diseases and infections to surrounding communities. This study aimed at determining the Health Effects of Chunga Dumpsite on surrounding communities in Lusaka, Zambia.

**Design and Methods:** This was a mixed method design and employed a cross sectional approach and case study conducted concurrently. Communities were stratified by distance into Stratum One (within 250 meters radius) and Two (above 250 to 500 meters). Quantitative data were collected from total 200 households thus 100 households from each stratum by using self-administered questionnaires. Qualitative data were collected from Two Key informants from the Local Authority and 15 participants using semi-structured interview guide. Both data were collected from December 22^nd^, 2018 to February 18^th^, 2019.

**Results:** Results revealed that smoke and various gasses emitted from burning and decomposing waste compromised quality of air in surrounding communities. Communities were infested with flies, mosquitoes and vermin. Consequently, (73.5%) complained of persistent coughing, (65.1%) suffered from malaria, (72.6%) complained of persistent headaches and (62.2%) had frequent diarrhoea cases in stratum one. Respiratory problems were more pronounced at night possibly due to high humidity which hindered pollutants to easily escape. However, these cases reduced drastically in stratum two.

**Conclusions:** Unsanitary dumpsites in LMICs including Zambia are commonly used as solid waste disposal facilities as opposed to sanitary landfills and become predisposing factors for spread of diseases. Short proximities to dumpsites by unplanned human settlements cause serious environmental challenges leading to public health risks to surrounding communities.

## Introduction

Dumpsites in low and middle income countries (LMICs) are commonly used for solid waste disposal as opposed to landfills. However, these facilities are generally ideal breeding grounds for disease transmitting agents such as mosquitoes, flies and rodents and therefore, they pose serious health issues to surrounding communities [1]. Consequently, open dumping of solid waste in many LMICs has been termed as a primitive stage of solid waste management because the systems applied lack scientific backing, are outdated and in-efficient [2, 3]. Therefore, these facilities create unsanitary environment and become predisposing factors for the spread of diseases and infections such as malaria, diarrhoea, cholera and tetanus [4].

The Chunga Dumpsite is the only government gazetted facility for solid waste (SW) disposal in Lusaka and is licensed to the Lusaka City Council (LCC) by the Zambia Environmental Management Agency (ZEMA) [5]. The facility was initially constructed to operate as a well-engineered commercial landfill. However, lack of adequate funding for many years, contributed to improper solid waste management. This led to deterioration of the conditions at the site and waste was dumped without compaction and soil cover [6, 11].

Municipal solid waste generation (MSWG) and disposal are among current global environmental and public health issues [7]. Human beings globally particularly in LMICs are faced with problem of the solid waste disposal as huge tonnage of waste is disposed in dumpsites which have negative health effects on surrounding communities [8]. Populations in fast growing cities in LIMCs are eminently at risk of being infected with diseases due to limited land to spare for proper disposal of waste, inadequate resources and lack of proper solid waste management systems [9, 10].

According to the Report of the Auditor General on Solid Waste Management in Zambia, the main methods of disposal of waste in major cities in Zambia including Lusaka, has been open dumping and incineration. Nevertheless, these methods of waste disposal have posed serious environmental and public health issues on surrounding communities that have evolved in the last decade as a result of rapid urbanization [11]. On the other hand, rapid urbanization in many LMICs has led to an increase in urban population growth resulting into increased waste generation per capita [12]. However, the increased footprint of waste generation has lagged behind the solid waste management systems and technology to those in the developed countries resulting in huge quantities of waste been deposited at dumpsites [13]

According to the 2010 Zambia National Census of Population and Housing, Zambia is one of the LMICs in sub-Saharan Africa that is highly urbanized and has about 40 percent of its inhabitants living in urban areas. Lusaka is the most urbanized city in the country with high population density and high population growth rate of 4.7 percent [14]. Consequently, the city has serious challenges with municipal solid waste management (MSWM) and has resulted in open burning and dumping of municipal solid waste (MSW) in illegal dumpsites [15]. The purpose of this study was to determine the Health Effects of Chunga Dumpsite on Surrounding Communities namely; Zanimhone West and Matero North in Lusaka, Zambia.

This study was guided by the conceptual framework developed by the researchers as shown in Fig.1 in which the concepts were adopted from the literature [16, 17].

**Fig. 1.**
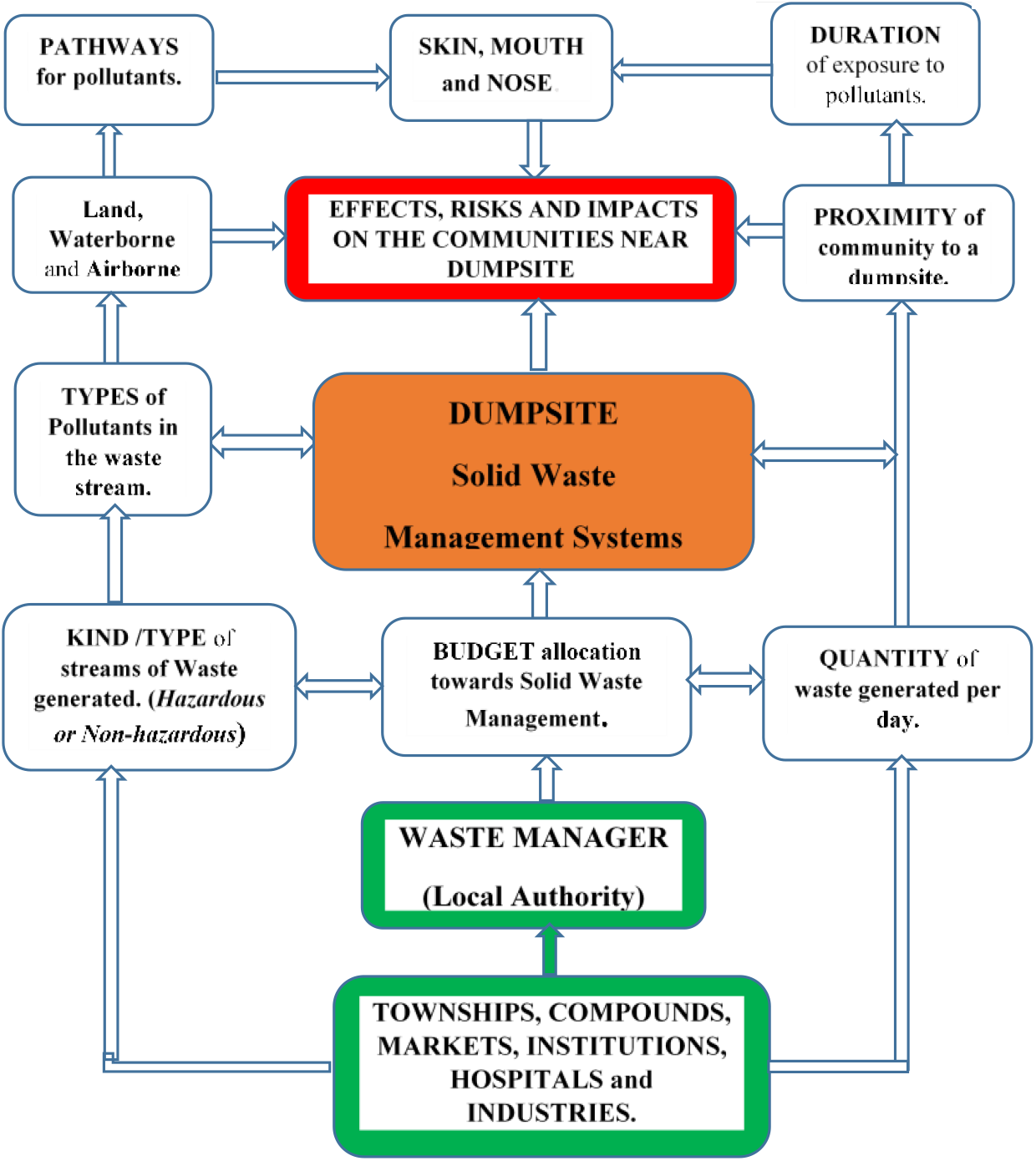
The conceptual framework that guided the study. **Source**: Researchers,(2018) adopted from (ISWA, 2015 and WHO, 2000).

## Materials and Methods

### Design and Study Site

This was a concurrent mixed method design where descriptive cross-sectional study and case study were conducted concurrently. For this study, participants included were those living within 0-500 meters proximities of Chunga Dumpsite in Lusaka. The study site is located on the outskirts of the northern part of Lusaka and positioned at 15°00’S and 28°02’E. According to Environment Protection Authority (EPA), Guidelines on Separation Distances on the establishment of polluting air emissions facilities for example a dumpsite, 500 meters from residential areas is recommended [18].

### Study Population and Sample Size

#### Quantitative arm

Non-probability sampling was employed throughout this study because the study communities were unplanned settlements and did not have a sampling frame. Listing of all housing units to create a sampling frame was not feasible due to limited time of the study.

The households of the communities from dumpsite were stratified by distance into two strata namely one (1) and two (2). All the households that were located within the radius of 250 metres were classified as stratum 1 and those above 250 metres to 500 metres as stratum 2. Thus, stratification of housing units by distance intended to create two groups of residents with similar socio-economic characteristics located at different proximities but both exposed to a common hazard (Chunga dumpsite). Purposive sampling methods were used to select identifiable and visible streets within 250 meters as well as households that were located above 250-500 metres. A total of 200 questionnaires were administered to two sets of household participants in stratum one and two.

#### Qualitative arm

Purposive maximum variation sampling methods were used to interview both male and female participants of different age groups, who were also exposed for different durations. The duration was the period a participant lived in the area. Hence, the nearest households to the dumpsite were sampled first and followed by those that were far from the dumpsite. For this study, 15 participants were interviewed despite reaching data saturation with 11 participants. The same procedure was repeated in stratum two. Further, these interviews engaged heads of households, male or female aged 18 years and above and who had lived in the study area for 3 months or more. Locations and distances of sampled households were determined and established by use of Global Positioning System (GPS). On the other hand, two key informants were purposively sampled from the City Council one from Waste Management Unit Office and another from Chunga Dumpsite Management Office.

### Technical information

#### Quantitative arm

Stata Version 14, (StataCorp. 2015. Stata Statistical Software: Release 14. College Station, TX: StataCorp LP.) was used to analyse quantitative data.

#### Qualitative arm

NVivo software version 12, (QSR International (1999) NVivo Qualitative Data Analysis Software) to manage qualitative data.

### Data Collection

Self-administered questionnaires with both open and closed ended questions were used to collect data for quantitative arm of the study. Heads of households either male or female aged 18 years and above who had lived in the area for at least three months and who gave their consent, participated in the study. All the interviews were conducted by the principal investigator and was assisted by qualified research assistant. Furthermore, the principal investigator conducted all observations at and around the dumpsite. A digital camera, audio recorders containing memory chips were used to collect and store data. English and a local language (Nyanja) were used for communication throughout data collection period.

### Ethical Considerations

Ethical approval was granted by the University of Zambia Bio-medical Research Ethics Committee (UNZABREC) and the National Health Research Authority (NHRA). Permission to conduct study at the dumpsite and surrounding communities was granted by the Lusaka City Council (LCC) who were the custodian of the facility. Detailed participant information sheet was availed to participants and only those who signed participant consent forms participated in the study. Privacy of participants was guaranteed by withholding identifiers such as names and addresses. Information collected was kept safely on computer with pass word only known to researchers.

### Statistical Analyses

#### Quantitative arm

Stata Version 14 was used to analyse data. Descriptive statistics were presented as frequencies and percentages using tables. Comparison of frequencies and percentages of health problems between households in stratum 1 and 2 was made to determine the extent to which the dumpsite effected the health of surrounding communities according to proximities.

#### Qualitative arm

NVIVO software version 12 was used to manage qualitative data. All audio recordings were safely stored on memory chips. All interviews were transcribed verbatim and data coding proceeded to analysis and interpretation. Some observed scenarios were presented in graphical form.

## Results

### Demographic Characteristics of the Participants

Table 1, presents data on background characteristics of the participants. Out of 200 participants, 70.5% were female and 29.5% comprised of males. Majority of the participants (51%) were within the economically active age group of between 26 to 40 years. However, the ages for the participants ranged from 18 to above 50 years. Meanwhile, majority (64%) were in informal employment and 42% of the participants had attained secondary education. Most participants (77.5%) were the owners of the houses who had lived in communities for more than 4 years.

**Table 1.** Socio- Economic and Demographic Data.

| Characteristic | Frequency | Percentage |
| --- | --- | --- |
| **Gender** |  |  |
| Male | 59 | 29.50 |
| Female | 141 | 70.50 |
| **Education** |  |  |
| None | 5 | 2.5 |
| Primary | 73 | 36.5 |
| Secondary | 84 | 42 |
| Tertiary | 38 | 19 |
| **Employment** |  |  |
| None | 12 | 6 |
| Informal | 128 | 64 |
| Public (formal) | 27 | 13.5 |
| Private | 11 | 5.5 |
| House wife | 22 | 11 |
| **Age** |  |  |
| 18 - 25 | 29 | 14.5 |
| 26 - 40 | 102 | 51.0 |
| 41 - 50 | 51 | 25.5 |
| Above 50 | 18 | 9.0 |
| **Home Ownership** |  |  |
| Land Lord | 155 | 77.50 |
| Tenant | 45 | 22.50 |
| **Water and Sanitation** |  |  |
| **Source of Water Supply** |  |  |
| Bore hole | 105 | 52.5 |
| Water Utility Companies (Chazanga Water and LWS) | 95 | 47.5 |
| **Type of Toilets** |  |  |
| Pit latrines | 124 | 62.0 |
| Flushable toilets | 76 | 38.0 |

### Proximities of surrounding communities to Chunga dumpsite and some reported health problems

Table 2, presents the proximities of the sampled households to the dumpsite in both strata. The reported symptoms included coughing, malaria, headaches, diarrhoea and cholera among others. Further, many households which, reported more health problems were located within 250 meters radius.

**Table 2.** Proximities of surrounding communities to Chunga dumpsite and some reported health problems.

| *Health Problem* | *Strat. 1 ( Within 250m)* |  | *Strat. 2 (251-500m)* |  | *Totals: Strat. 1 and 2* |  |  |
| --- | --- | --- | --- | --- | --- | --- | --- |
|  | **Female**<br>**n (%)** | **Male**<br>**n (%)** | **Female**<br>**n (%)** | **Male**<br>**n (%)** | **0 – 250m:**<br>**Strat 1 n (%)** | **251–500m:**<br>**Strat 2 n (%)** | **Total**<br>**n (%)** |
| *Coughing* | 67 (50.8) | 30 (22.7) | 27 (20.5) | 8 (6.1) | 97 (73.5) | 35 (26.5) | 132 (100) |
| *Headaches* | 56 (49.5) | 26 (23) | 23 (20.4) | 8 (7.1) | 82 (72.5) | 31 (27.5) | 113 (100) |
| *Malaria* | 48 (44) | 23 (21.1) | 32 (29.4) | 6 (5.5) | 71 (65.1) | 38 (34.9) | 109 (100) |
| *Diarrhoea* | 52 (46.8) | 17 (15.3) | 29 (26.1) | 13 (11.7) | 69 (62.2) | 42 (37.8) | 111 (100) |
| *Cholera* | 4 (50) | 0 (0) | 4 (50) | 0 (0) | 4 (50) | 4 (50) | 8 (100) |

### Practices of Solid Waste Management at household level

As presented in Table 3, the results showed that the majority (73.5%) of the participants did not dispose waste at the dumpsite as opposed to a small percentage (26.6%). The majority (93%) in both strata used polyester sacks as waste receptacles for temporal storage of waste instead of containers covered with lids. The majority (48%) from stratum one were aware of health problems that may have been associated with poor SWM.

**Table 3:** Practices of Solid Waste Management (SWM) at household level.

| Practices of SWM<br>(Household level) | Strat. 1 (0 – 250m) |  | Strat. 2 (251-500m) |  | Strat. 1 and 2 |  |  |
| --- | --- | --- | --- | --- | --- | --- | --- |
|  | Female (%) | Male (%) | Female (%) | Male (%) | 0 – 250m Strat 1(%) | 251–500m Strat 2(%) | Total (%) |
| Waste disposed at Chunga dumpsite | 31 (58.5) | 12 (22.6) | 9 (17.0) | 1 (1.9) | 43 (81.1) | 10 (18.9) | 53 (100) |
| Waste disposed through other methods | 38 (25.9) | 19 (12.9) | 63 (42.9) | 27 (18.4) | 57 (38.8) | 90 (61.2) | 147 (100) |
| **Types of Waste Bins for storage** |  |  |  |  |  |  |  |
| Recommended containers covered with lids | 6 (42.9) | 0 (0) | 5 (35.7) | 3 (21.4) | 6 (42.9) | 8 (57.1) | 14 (100) |
| Polyester sacks | 63 (33.9) | 31 (16.7) | 67 (36.0) | 25 (13.4) | 94 (50.5) | 92 (49.5) | 186 (100) |
| **Knowledge about Health Problems due to poor SWM** |  |  |  |  |  |  |  |
| Not able to mention at more than 3 | 3 (16.7) | 1 (5.6) | 13 (72.2) | 1(5.6) | 4 (22.2) | 14 (77.8) | 18 (100) |
| Able to mention at least 3 | 26 (25.7) | 16 (15.9) | 37 (36.6) | 17 (16.8) | 42 (41.6) | 59 (58.4) | 101 (100) |
| Able to mention at more than 3 | 40 (46.5) | 14 (16.3) | 22 (25.6) | 10 (11.6) | 54 (62.8) | 32 (37.2) | 86 (100) |

### Participants’ views about location and closure of the Chunga Dumpsite

As indicated in Table 4, majority of the participants (87%) believed that the location of the dumpsite had negatively impacted on the quality of their environment and health. They mainly complained of the plume smoke and bad odour that emanated from the dumpsite and suggested for the relocation of the facility. However, a smaller percentage of participants (3%) maintained that the dumpsite had no impact on their health and there was no need to relocate it since it was of economic benefit for their livelihood.

**Table 4:**
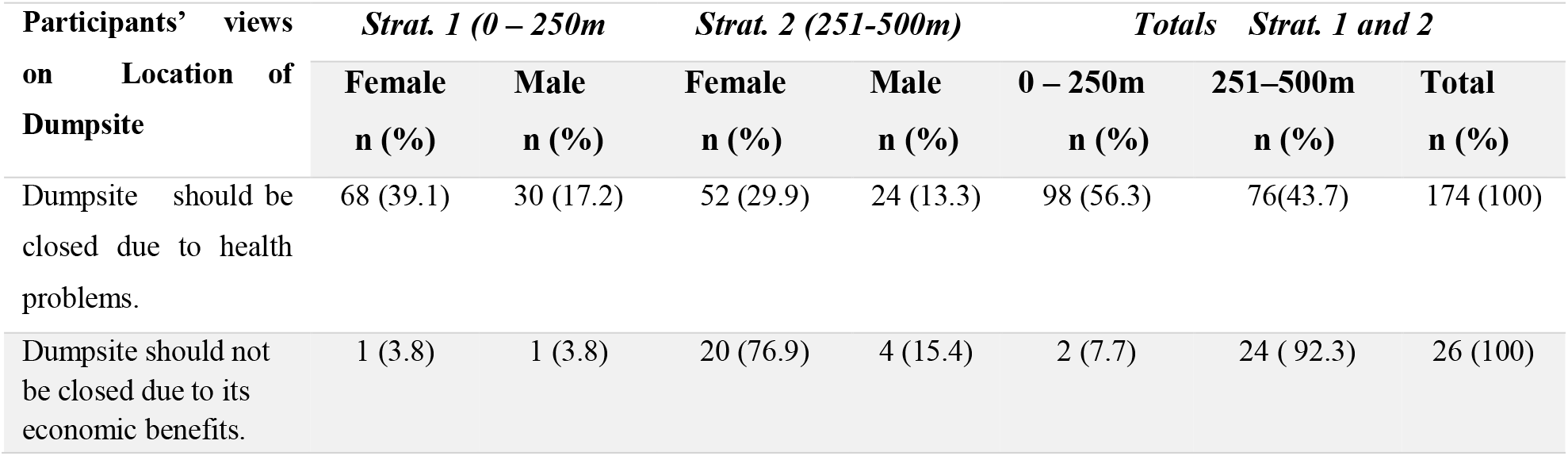
Participants’ views about location and closure of the Chunga Dumpsite.

### Perceived Environmental and Health Effects of Chunga Dumpsite by Participants

The participants reported that some mixed wastes were deposited at the dumpsite including municipal, healthcare, industrial and electronic wastes. These were confirmed through observations. The site was observed to be filthy with swarms of flies everywhere, dusty including plume smoke from burning waste and the site was smelly. Thus, following themes and their related quotes describe the perceived environmental and health effects of the dumpsite.

### Air Pollution

Air pollution was one of overarching themes with subthemes that included bad odour, smoke, and dust. The following subthemes explain the participant narration of the pollutants:

#### Bad Odour

Bad odour was one of many issues which participants complained about. The following quote by a participant explains:

> *“…concerning the waste that is dumped here, it is a very serious issue to talk about. As can be seen, there are many flies due to different types of biodegradable waste that is dumped which exude bad odour. Among the things dumped along with waste are stillborn babies, rotten skins of animals from abattoirs and condemned food stuffs from chain stores. In some cases, refuse trucks get marooned around dumpsite and exuding bad stench …”* **(Participant No. 03)**

It was further observed that refuse trucks (Fig.2) were packed without offloading the waste for two days due lack of heavy duty machinery to level waste.

**Fig. 2.**
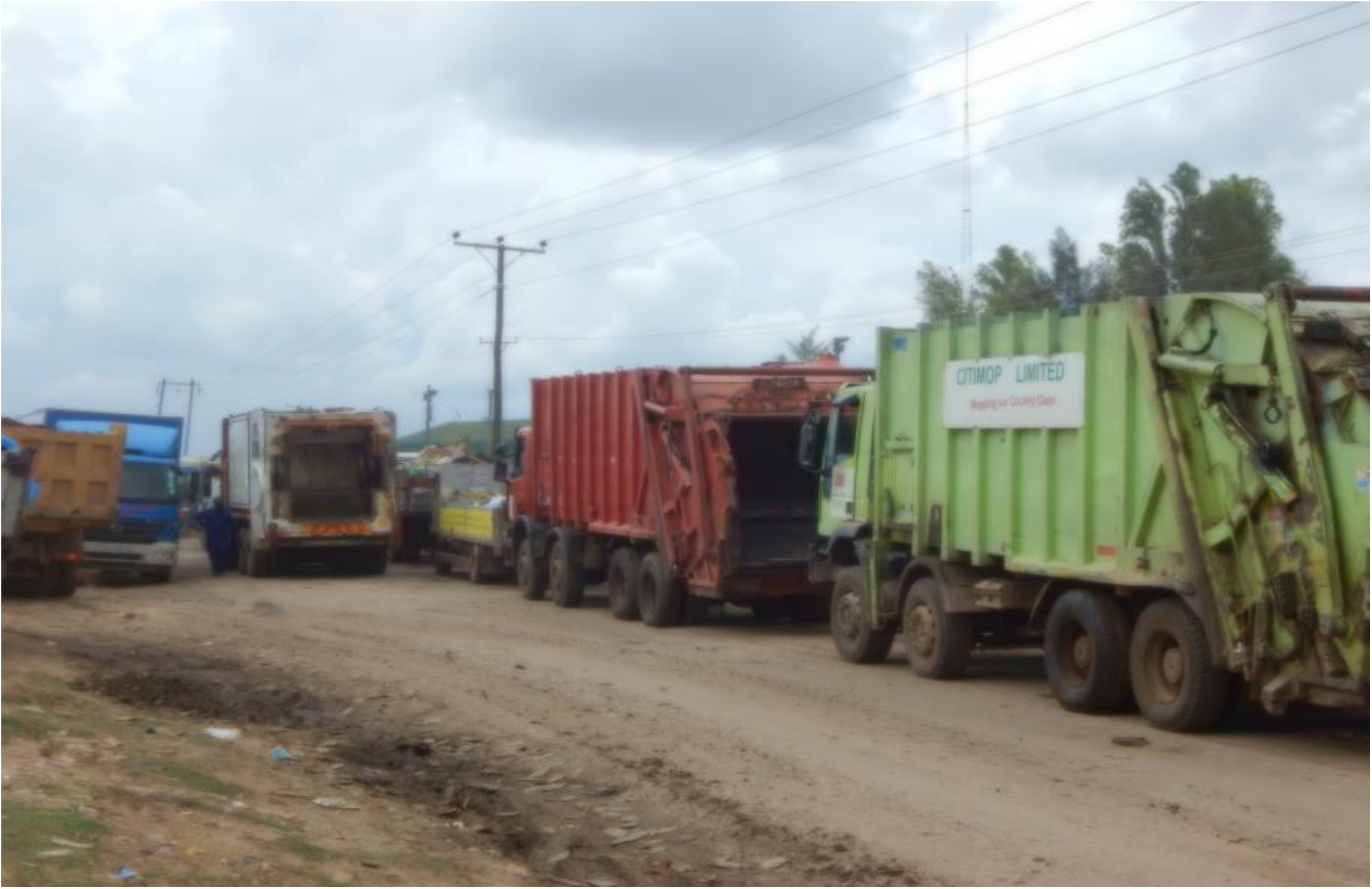
Loaded refuse trucks near houses marooned at the dumpsite.

#### Plume Smoke

The smoke that emanated from burning wastes was reported to have polluted and compromised the quality of air in the communities around dumpsite as explained by the key informant at the dumpsite:

> **“***…*.*the potential health hazards at this site are many. Fires burn continuously and get worse in the dry season when thick smoke is produced. Consequently, the smoke from the fires are health risks to our people around here. The worst affected are those who live across the dumpsite where wind normally blows. As a result, many people especially the asthmatic patients bring complaints to our office every day about been choked with smoke leading to persistent coughing “***(Key Informant 01)**

Further, the researchers observed and captured on camera pockets of areas burning and smoke circulated continuously (Fig. 3). Meanwhile, scavengers who looted the area for valuable items ignited the fires but site managers could not control the situation due to inadequate security around site perimeter.

**Fig. 3.**
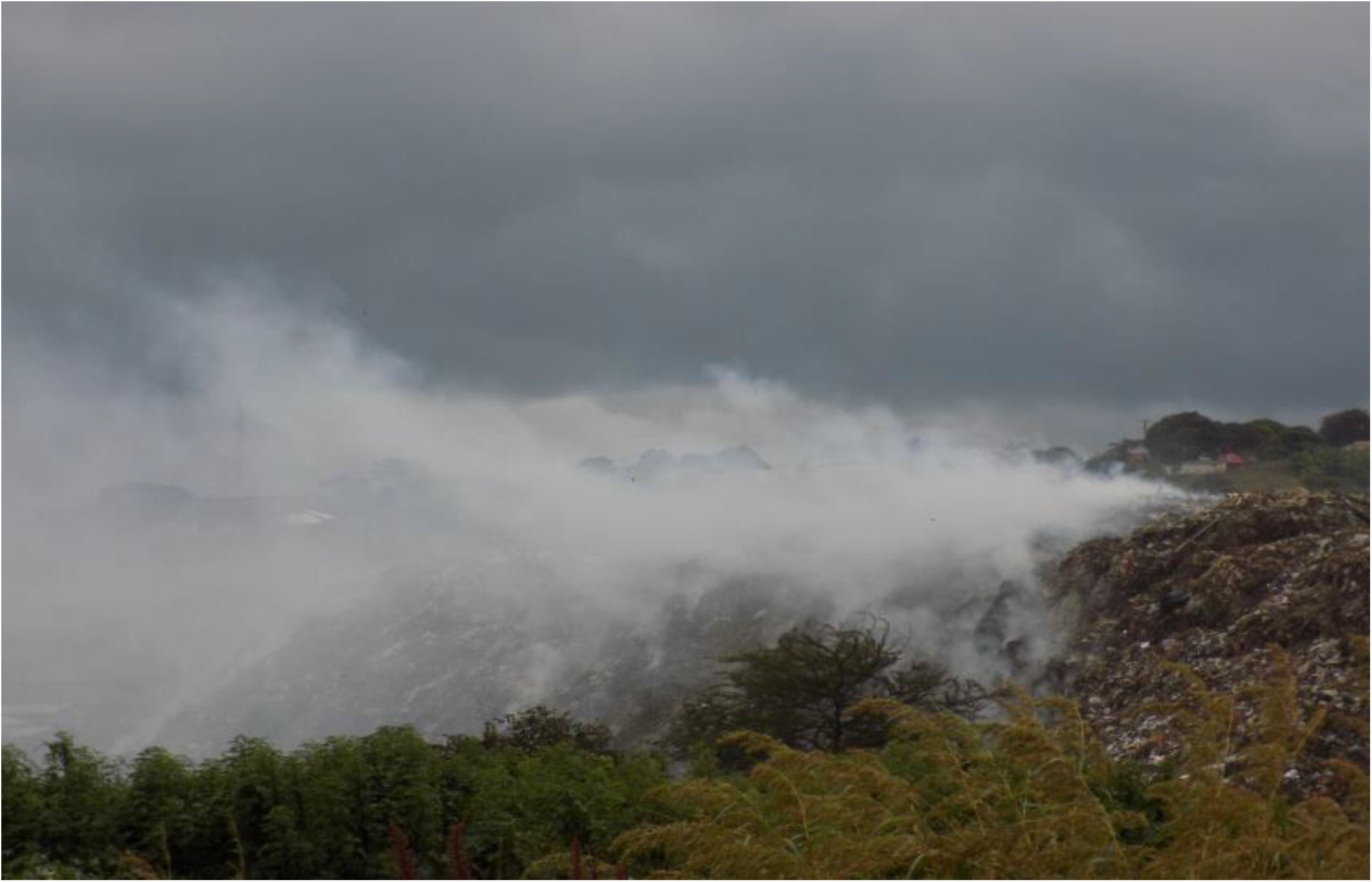
Plume smoke from burning waste at the dumpsite.

### Challenges of unsanitary Solid Waste Disposal faced by the Lusaka City Council (LCC)

This study explored the challenges of unsanitary solid waste disposal faced by the LCC. Interviews with key informants were supplemented by the observation notes by the researchers.

#### Lack of heavy-duty machinery by LCC at the Dumpsite

The key informant at the dumpsite said that the facility lacked functional heavy-duty machinery to level, compact and cover waste with soil according to schedules. Therefore, waste was absurdly dumped without compaction leading to creation of an open dumpsite. The following was a response by one of the key informants:

> “*The initial design of the site was a well-engineered land fill and was constructed from 2003 to 2005 with the help of the Danish Government. The landfill was later handed over to the Lusaka City Council (LCC) but along the way LCC failed to manage it properly due to inadequate funding, lack of skilled manpower to work on specialised pieces of equipment as well as lack of political will*. **(Key informant No. 01)**

## Discussions

This study sought to determine the health effects of Chunga Dumpsite on the surrounding communities in Lusaka, Zambia. Thus, the main focus was on the health effects of the dumpsite on surrounding communities in relation to the proximities of households to dumpsite.

The age limit for most of the participants was between 26 to 40 years. This rightly represented the country’s population and workforce which is youthful. There was also a higher participation of women in the study and this too represents the country’s gender distribution as highlighted in the results of the 2010 Zambia National Census of Population and Housing which showed that women were more than men [14]. Additionally, most participants who resided around dumpsite area owned most of the housing unities and had lived there for more than four years.

Further, these people were mainly self-employed with a monthly income of less than US$150. Majority of them had attained primary and secondary education and this is in line with the government’s policy on Universal Free Primary Education Policy which envisioned to curb illiteracy, inaccessibility, unaffordability, inequality and high dropout rates in school [19, 20]. Hence, it is assumed that this population group is literate enough to respond positively to health education campaigns on prevention of disease outbreaks and respecting the international guidelines on safe distance to the dumpsite [21].

However, the results of this study showed a disregard for such guidelines because 50% of the participants lived within 0-250m radius. Based on documents assessed for this dumpsite, the study revealed that the facility was initially situated on the outskirts of the city but its surroundings became encroached as city population grew. Consequently, some households have been established as close as three meters away from dumpsite which is against the international guidelines on Separation Distances for Airborne Emissions facilities like a dumpsite [18]. Meanwhile, exposure to continuous plume smoke from burning waste and pungent smells of decomposing organic matter is associated with poor air quality which compromises human health [24, 25].

Consequently, comparison of health related problems between the households in stratum 1 (0-250m) and those in stratum 2 (251-500m) was made. Most of participants who lived in stratum 1 perceived that the dumpsite had negatively impacted on quality of air and the environment. Hence, they suspected that most the illnesses and symptoms like headaches, coughing, malaria and diarrhoea were attributed to the presence of the dumpsite.

Participants from stratum1 confirmed that their households were too close to the dumpsite and viewed the location as a threat to their health. As observed, the residents cited plume smoke, bad odour, and dust including flies and mosquitoes as their main challenges to their health. Therefore, they proposed for relocation of dumpsite to avert further negative impacts on their health and environment. Thus, Salam (2010) in his research at the Golf Course dumpsite in Manzini City observed similar results [26].

Meanwhile, households that were closer to the dumpsite (stratum 1) reported more different health problems. This observation showed that the likelihood of a person living in this location to complain of symptoms was higher than one who lived in stratum 2. Hence, this is in tandem with some reports by some researchers that as distances increased from dumpsite as an exposure site, incidences of illness reduced drastically [22, 23].

In this regard, the risks of disease outbreaks for communities around the Chunga dumpsite is higher due to poor solid waste management. Thus, the risks are higher for households in stratum1 due to possible higher concentrations of various poisonous gasses like carbon monoxide from subsurface combustion (anaerobic) and carbon dioxide from surface combustion (aerobic) of waste including uncontrolled release of methane from anaerobic decomposition of waste [22]. Hence, too much inhalation of these gasses could have led to health problems like coughing, headaches and fatigue by residents in the nearby communities [23]. Further, communities whose proximities are close to dumpsite risk some disease transmitting agents to easily invade households and contaminate utensils and food which if consumed may cause various illnesses [24].

However, this could have also been attributed to poor solid waste management at household level. For this study, inappropriate disposal of waste by participants at household level was rampant. For example, 92% used polyester sacks as primary waste storage containers which leaked leachate into the environment and attracted flies. This often risks environmental contamination which may manifest through contamination of both surface and ground water. The, open dumping of waste at household level provided favourable conditions and served as ideal breeding grounds for disease transmit agents. These agents may transmit pathogens either from the waste dump or other similar sources into surrounding communities [25].

Meanwhile, it was interesting to note that cases of cholera were insignificant in both strata despite the backdrop of the city recording confirmed spikes of Cholera cases in the city. For this study, only 4% of participants reported possible cases of Cholera in each stratum. Meanwhile, other studies in Sierra-Leone found that communities which lived near the dumpsite were usually victims of cholera outbreaks (Sankoh *et al*., 2013). The low cholera positivity in this study could have been attributed to the cholera health campaign messages by the Ministry of Health (MoH) during the same period the study was conducted. This could also be attributed to improved water and sanitation facilities that were observed in the communities.

On the other hand, one of the Key informants at the dumpsite confirmed that, lack of heavy-duty machinery at the dumpsite had hindered sustainable waste management. The huge tonnage of waste that was discarded at the facility was not regularly levelled to the standard thickness of three metres, and each layer of waste was not compacted and covered with soil of not less than 30 centimetres before another layer of waste could be spread over [9].

Therefore, some waste become airborne and scattered into surrounding communities during windy periods thereby compromising the aesthetic values of the communities [27]. On the other hand, the situation exacerbated surface fires which produced plume smoke and polluted air in nearby communities through the process of diffusion particularly when humidity became high and when wind blew in specific directions. These findings were earlier highlighted in 2007 in the Report of the Auditor General on Solid Waste Management in Zambia, which emanated from its Environmental Audit on Waste Management [9].

## Conclusions

Unsanitary dumpsites in LMICs including Zambia are commonly used as solid waste disposal facilities as opposed to sanitary landfills. These, in the process became ideal breeding grounds for disease transmitting agents and sources of exposure to pollutants. Short proximities to dumpsites by unplanned human settlements cause serious environmental challenges leading to public health risks to surrounding communities. Hence, disease outbreaks were common in such settings where poor solid waste management manifested with close proximities to human settlements. For this study, percentages of health problems associated with effects of dumpsite, decreased drastically with increased distances from the exposure site.

Despite some limitations, confounding factors and effect modifiers, these findings may indicate the real health risk factors that are existing among the surrounding communities around Chunga dumpsite.

Therefore, the council should improve solid waste management at the site through improved budgets towards solid waste management. Communities should be sensitised about the risks of exposure to pollutants from dumpsite to avert further negative impacts on public health, environment and aesthetic values. Thus, the dumpsite should be closed gradually whilst a new landfill is constructed on the outskirts of the city. Further studies to establish the quality of air inhaled and quality of water consumed from the boreholes are recommended.

## Data Availability

Data set for this study was generated and is in the custody of the researchers that can be accessed upon request.

## Data availability

Data set for this study was generated and is in the custody of the researchers that can be accessed upon request. Further, references are provided for other secondary data used in this study.

## Authors’ contributions

Wilson Chibwe (WC) took up the responsibility of designing the study, data collection, conducting the literature review and data analysis. Dr. Alice Ngoma Hazemba (ANH) was responsible for structuring the results section and to prepare the manuscript as well as editing. Aroan Mbewe (AM) was involved in data analysis and provided some necessary revisions for the manuscript preparation.

## Acknowledgements

The authors wish to thank all the participants for their wonderful cooperation and their contributions that have made this research a success.

